# The limits of estimating COVID-19 intervention effects using Bayesian models

**DOI:** 10.1101/2020.08.14.20175240

**Authors:** Patrick Bryant, Arne Elofsson

**Affiliations:** Science for Life Laboratory and Department of Biochemistry and Biophysics, Stockholm University, Box 1031, 17172 Solna, Sweden

## Abstract

To limit the rapid spread of COVID-19, most governments have introduced different non-pharmaceutical interventions, which might have severe costs for society. Therefore, it is crucial to evaluate the most cost-effective interventions, using, for instance, Bayesian modelling. Such modelling efforts have deemed lockdown to account for 81% of the reduction in R_0_, contributing to government policies. Here, we show that these conclusions are unsupported and that policies therefore should not be based on these studies.

*https://www.eurosurveillance.org/for-authors*

## Background

Due to the rapid spread of COVID-19 across Europe, and the quick increase in cases and deaths in some countries, European countries introduced non-pharmaceutical interventions to limit virus spread^1^. Efforts to elucidate the impact of interventions through computational modelling have been made^2–4^. Still the actual effects of these interventions are hard to differentiate from each other, as many interventions were introduced almost simultaneously. This communication focuses mainly on the deficiencies with a recent Bayesian model used to estimate the effect of five different interventions across 11 European countries^2^. Still, it is also relevant for other modelling attempts, including our own, where we try to use mobility in various sectors of society to estimate their effects^5^.

A recent study^2^ uses MCMC simulations^6,7^ to infer impacts on the basic reproductive number (R_0_) through governmental non-pharmaceutical interventions (NPIs). From the estimated R_0_, the number of daily cases is predicted, which then through an infection-to-death distribution, is used to infer the number of daily deaths. The model fits the number of daily deaths to the observed number of deaths^8^ using a Bayesian posterior distribution. The paper reports that current interventions have been sufficient to drive R_0_ below 1 (which is necessary for the number of infected people to decrease) for all 11 European considered countries. They *“show that major non-pharmaceutical interventions and lockdown, in particular, have had a large effect on reducing transmission”*. The authors conclude that lockdown accounts for 81% [75% – 87%] of the reduction in R_0_, i.e. that all other interventions only have minimal effects on the decline of R_0_.

## The issue

The peculiar aspect of the claim that lockdown accounts for 81% of the reduction in R_0_, is that Sweden did not implement any lockdown, but still see a similar decrease in R_0_ as the other countries, even though the other NPIs were reported to have no substantial effect on R_0_. To solve this problem, as compared with the authors’ earlier work^9^, which showed a significantly higher R_0_ for Sweden, they invoke a country-specific last intervention parameter, which is only implemented for Sweden^10^ (see equation *i*). The “last intervention” parameter is multiplied with R_0_, and can therefore be seen as a parameter adjusting the model for Sweden independently. As can be seen, when analysing the posterior distributions of the intervention parameters, the “last intervention” parameter for Sweden results in 73.5 % of Sweden’s reduction in R_0_ (Figure 1). The last intervention impact on R_0_ is not reported or discussed in the Nature publication, possibly misleading decision-makers on the importance of lockdowns.

**Figure 1.**
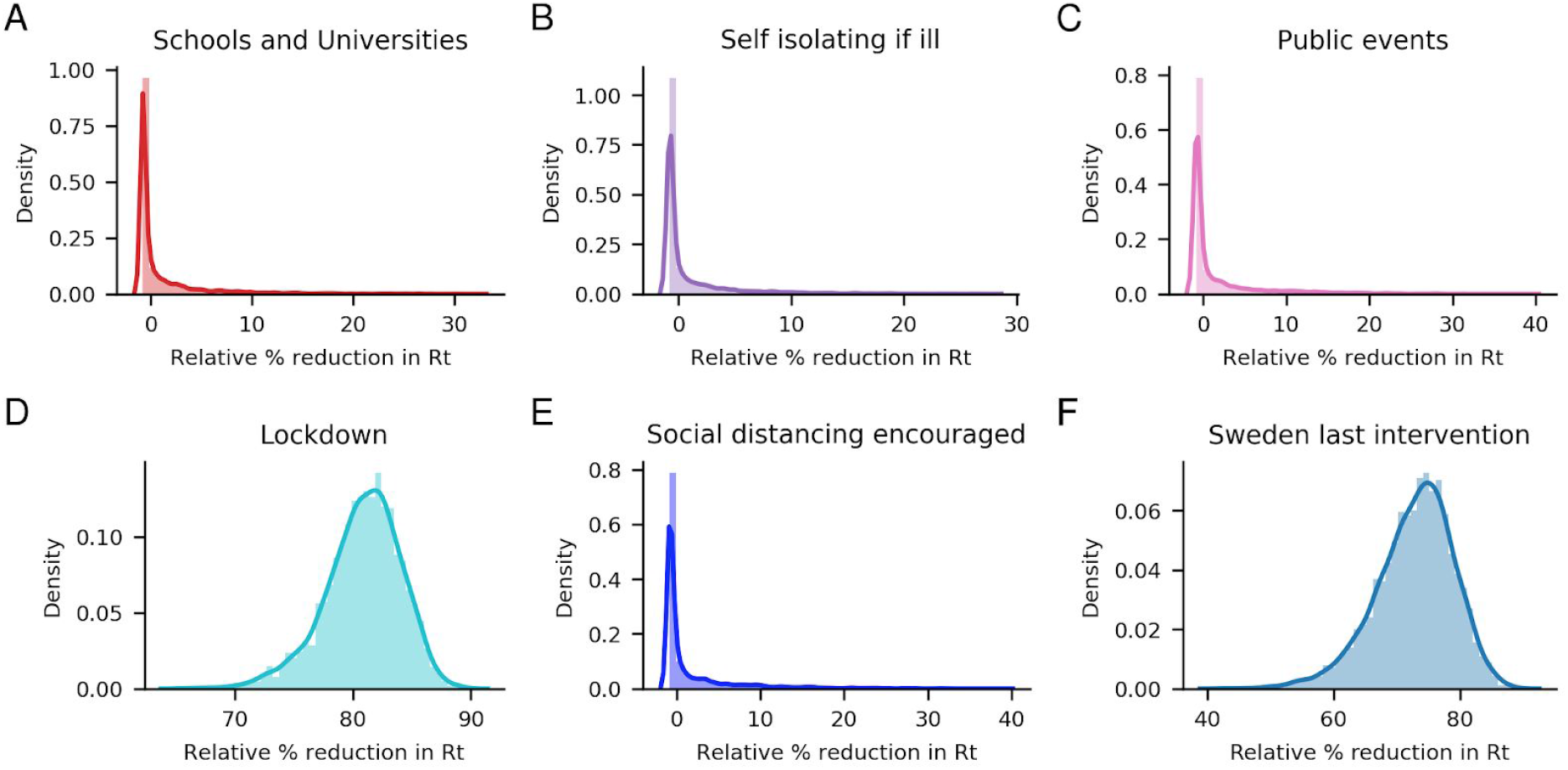
Posterior distributions of relative reduction in R_0_ for each intervention and the “last intervention” parameter only introduced for Sweden. The median reductions are –0.7%, –0.7%, –0.7%, 81.1% and –0.6 % for NPIs Schools and Universities, Self-isolating if ill, Public events, Lockdown and Social distancing encouraged respectively. The last intervention, only implemented for Sweden, has a median reduction of 73.5%.

This is the equation used for estimating the impact of NPIs on R_0_ in the MCMC simulations

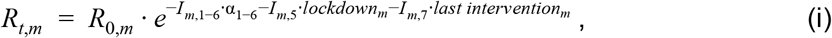

where I_m, 1–7_ states if country m has introduced the following interventions, on the given day, (1) closing of schools and universities, (2) Self-isolating if ill, (3) banning of public events, (4) the first intervention introduced, (5) lockdown, (6) social distancing encouraged and (7) last intervention. The additional country-specific parameters added for the lockdown (here referred to as “extra lockdown”) and last intervention are sampled from the following distributions:

lockdown ∼ normal(0,gamma)

last intervention∼ normal(0,gamma)

gamma ∼ normal(0,.2)

To analyze the importance of the last intervention option only introduced for Sweden, we remove it (see equation *ii*). The resulting reduction in R_0_ for Sweden is identical within statistical margins, but the fit to the daily deaths is worse (Figure 2). Interestingly, the banning of Public events (Sweden’s last introduced NPI) now becomes more important (median reduction of 50.1%), being almost on par with that of the lockdown (median reduction of 64.9%, Figure 3).

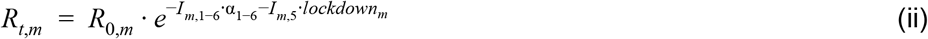

**Figure 2.**
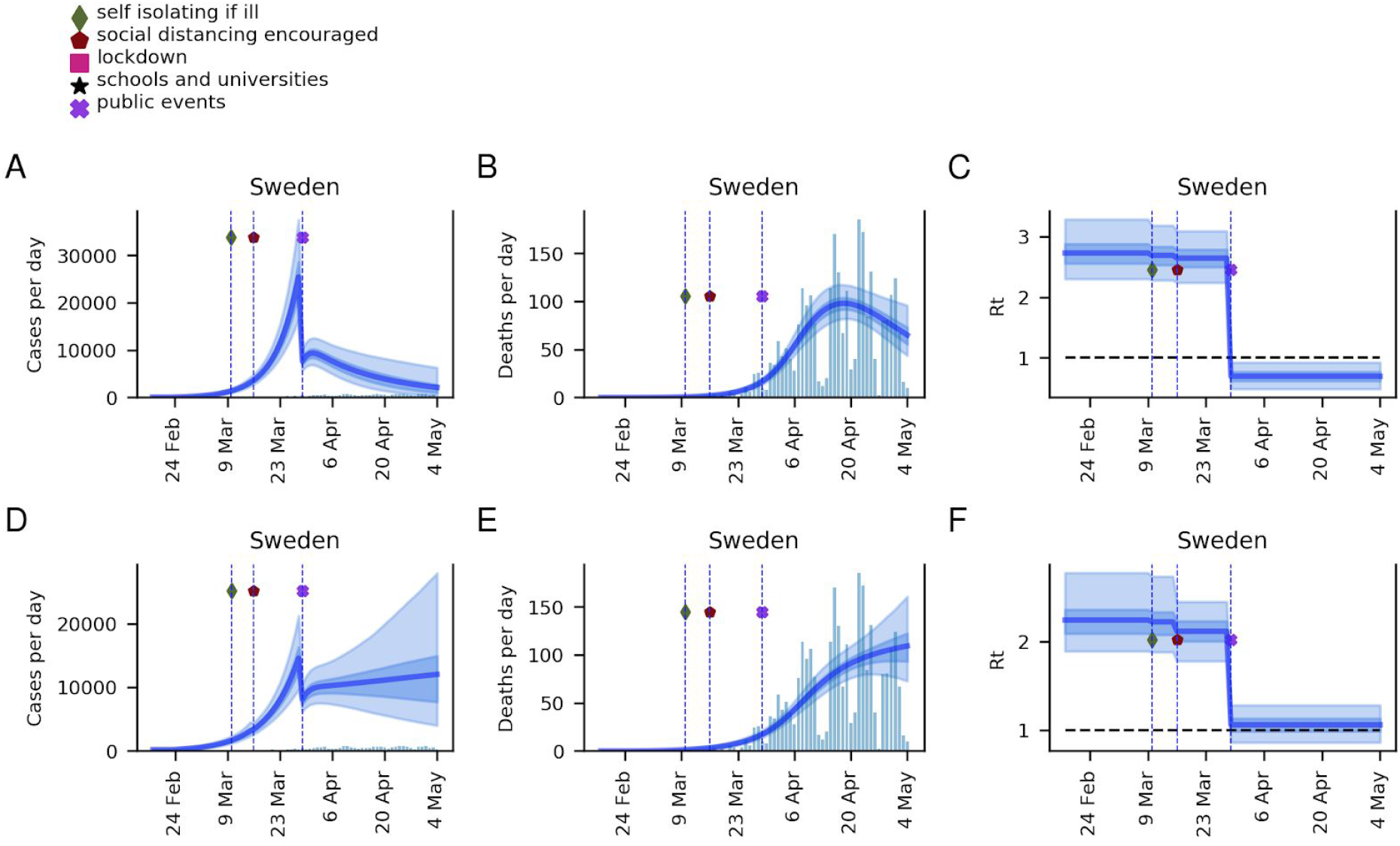
Modelling results for Sweden with (A, B, C) and without (D, E, F) the introduction of the “last intervention” parameter. The thick blue lines correspond to the means, while the darker and lighter shades of blue correspond to the 50 and 95 % confidence intervals respectively.

**Figure 3.**
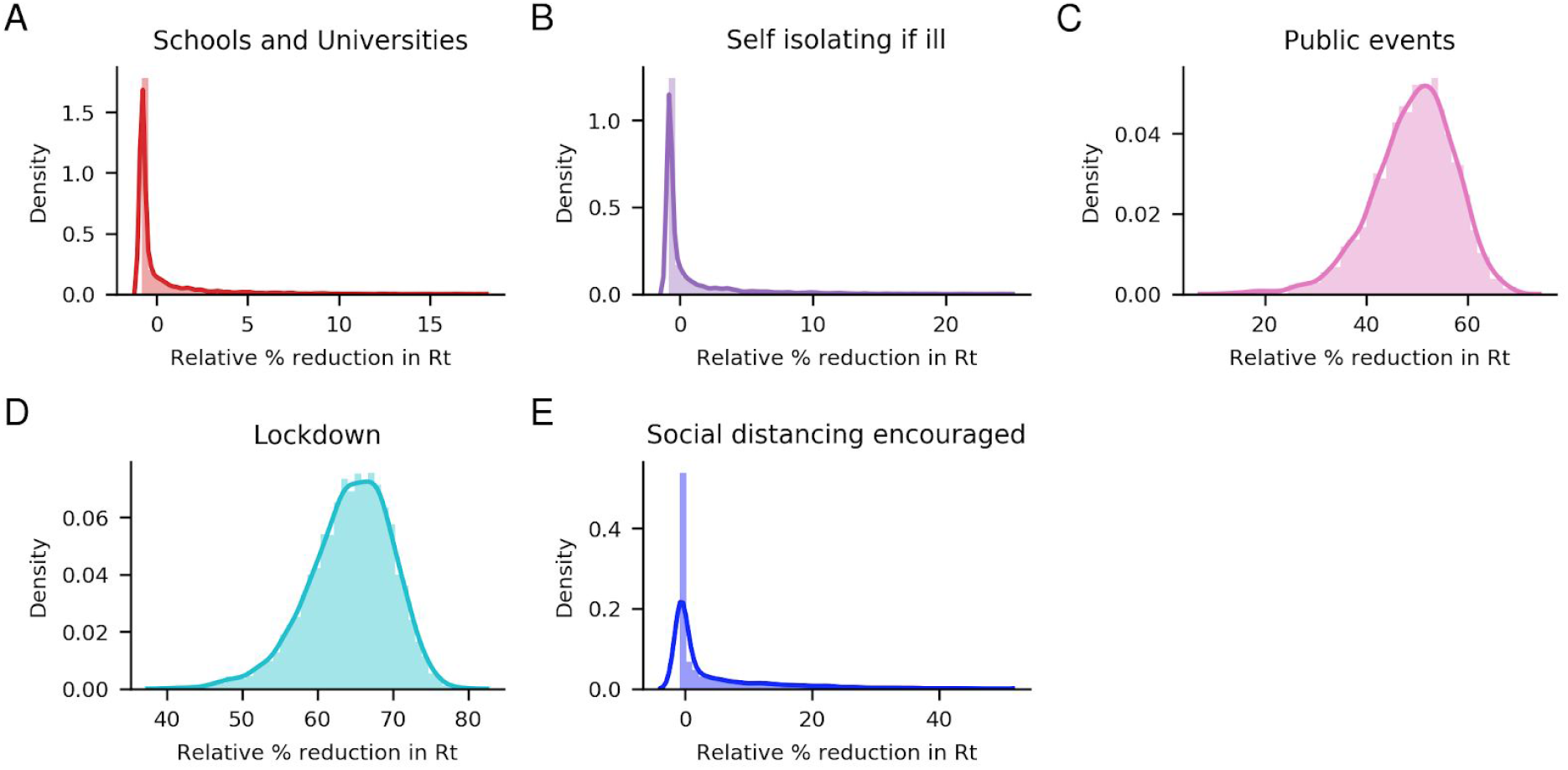
Posterior distributions of relative % reduction in R_0_ if the last intervention option only introduced for Sweden is removed. The median reductions are –0.7%, –0.7%, 50.1%, 64.9% and –0.1 % for NPIs Schools and Universities, Self-isolating if ill, Public events, Lockdown and Social distancing encouraged respectively.

Finally, suppose the “extra lockdown” introduced for all countries except for Sweden and the “last intervention” options only added for Sweden are removed (equation *iii*). In that case, the banning of public events now becomes the NPI with the most importance (reduction of 78%). Now also the effect of the closing of schools and universities, and social distancing rise (reductions of 5.4%, and 5.3% respectively), while the self-isolating if ill importance remains unchanged (median 0.7 %), and the lockdown importance greatly diminishes (0.2 %), Figure 4.

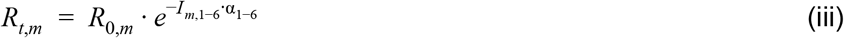

**Figure 4.**
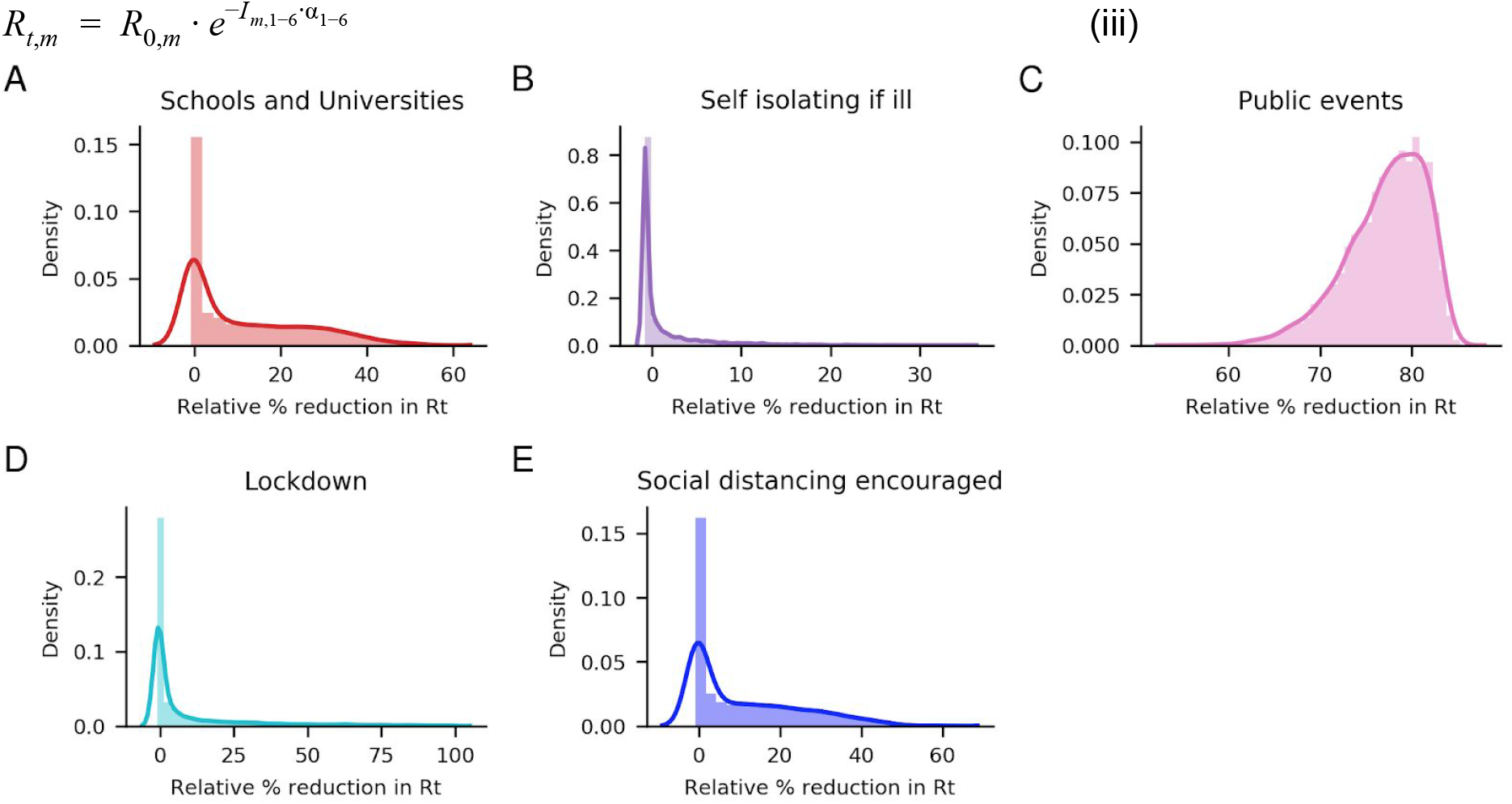
Posterior distributions of relative % reduction in R_0_ if the extra lockdown option and the last intervention option only introduced for Sweden is removed (see equation iii). The median decreases are 5.4%, –0.7%, 77.9&, 0.2% and 5.3 % for NPIs Schools and Universities, Self-isolating if ill, Public events, Lockdown and Social distancing encouraged respectively.

## Discussion and Conclusion

The Bayesian model used by ICL to estimate the impact of each intervention appears to be sensitive to small perturbations. The reason the lockdown seems to have such high importance is likely due to it being the last intervention implemented, and not because it had the most substantial impact. We highlight this by showing that the importance of the “last intervention” diminishes when treating Sweden in the same way as all other countries.

Further, the modification of the initial model^9^ enables adjustments to the daily deaths from the model in each country. It thus creates the possibility of fitting a wide range of data to the observations but loses the ability to identify cross-country patterns. We believe that allowing country-specific effects of the NPIs makes it very difficult to estimate the relative impacts of NPIs, since there is no possibility of distinguishing between NPI effects and other country-specific effects, e.g. population density.

In conclusion, it is peculiar that the model displays an almost identical change in R_0_ in all countries, dropping sharply below one at the final NPI, independently on the nature of that NPI. In reality, all countries had different NPIs implemented at different time points, likely with varying strength and efficiency, and it is quite likely that NPIs such as enforcing social distancing at least had some effects, not seen in the models. Given the importance the initial report had on government policies and the fact that we show here that the conclusions made about the significance of the lockdown are not entirely correct, we do think that we should pinpoint this to readers and policymakers. Correct assumptions on the effects of NPIs are becoming even more urgent as many nations still are imposing different NPIs, and that these might go on for an extended period (https://www.bbc.com/news/explainers-53640249, last accessed 20200811).

## Data Availability

All data and code is available from https://github.com/patrickbryant1/COVID19.github.io/tree/master/ICL_critique

## Acknowledgements

We acknowledge Claudio Bassot’s contribution by sharing the Imperial College. Without this information, this study would not be possible. We are also grateful to various colleagues and friends that contributed to the discussion. Finally, we thank the authors of the Imperial College London report for making their data and model freely available.

